# Difference in presentation, outcomes, and hospital epidemiologic trend of COVID-19 among first, second, and third waves in Dhaka Medical College

**DOI:** 10.1101/2022.12.14.22283379

**Authors:** Reaz Mahmud, Md. Ashikul Islam, Md. Emdadul Haque, Md. Dewan Azmal Hussain, Mohammad Rafiqul Islam, Farhana Binte Monayem, Mostofa Kamal, Hashmi Sina, Mohammad Fakhrul Islam, Ponkaj Kanti Datta, S.K. Jakaria Been Sayeed, Sabbir Ahmed Dhali, Khairul Islam, Rifat Hossain Ratul, SK Md. Rubaed Hossain, Habib Naziat Prince, Ahmed Hossain Chowdhury, Kazi Gias Uddin Ahmed, Md.Titu Miah, Md. Mujibur Rahman

**Affiliations:** Department of Neurology, Dhaka Medical College. Dhaka, Bangladesh; National Institute of neurosciences and Hospital. Dhaka, Bangladesh; National Institute of Chest diseases and Hospital; Department of Medicine, shaheed Suhrawardy Medical college; Sarkari karmachari Hospital. Dhaka, Bangladesh; Department of Medicine, Dhaka Medical College; Department of Medicine, Dhaka Medical College, Dhaka, Bangladesh; COVID-19 post-acute care and follow-up clinic, Dhaka Medical College; North South University; Department of Medicine, Bangabandhu Sheikh Mujib Medical University, Dhaka. Bangladesh

## Abstract

**Background:** This study aimed to examine the differences in epidemiologic and disease aspects among patients with COVID-19

**Methods:** We reviewed the hospital records between April 2020 and September 2021 and followed up on the patients for post-COVID complications.

**Findings:** Older adult patients were predominantly affected during the first and second waves, followed by middle-aged patients. Men were predominantly admitted, considering the three waves; although more women were admitted in the second wave. Cough was more common in the second and third waves than in the first wave 522 (59.7%). Respiratory distress was the most common in the third wave, 251(67.1%), and least common in the first wave 403 (46.1%). Anosmia was more common in the third wave 116 (31.2%). In the third wave, patients presenting in a critical state 23 (6.2%) and severe disease 152 (40.8%) were more common. The hospital admission median (IQR) was longer in the first wave, 12 (8–20), than in other waves. More patients were admitted in the first wave (52%) than in the other waves, and patients received more oxygen in the third wave (75%) than in the other waves. Death occurred more commonly in the first wave (51%) than in the other waves. Patients were investigated more commonly in the first and third waves than in the second wave. The positivity rate was high in the third wave (22.8%) than in other waves. In the third wave, the positivity rate was higher in women (24.3%) than in men. Post-covid cough increased in the second wave and fatigue was higher in the third wave than in other waves. Tiredness and memory loss was greater during the second wave than in other waves.

**Conclusion:** This study revealed that the presenting symptoms, outcomes, and epidemiologic trends differed during the COVID-19 waves.

## Introduction

On December 31 World Health Organization (WHO) formally reported a case of atypical pneumonia in Wuhan City, China [1], which was later named coronavirus disease-19 (COVID-19) caused by severe acute respiratory syndrome coronavirus 2 (SARS-CoV-2) [2]. On March 11, 2020, WHO declared the novel coronavirus (COVID-19) outbreak a global pandemic [3].

The WHO classified the phases of the pandemic as phases 1–3(predominantly animal infection, few human infections), phase 4(sustained human-to-human infection), phases 5 and 6 (widespread human infection), post-peak (possibility of recurrent events), and post-pandemic (disease activity at seasonal level) [4]. Most of the worst pandemics lasted for several years [5]. After studying the transmission dynamics of SARS-CoV-2, researchers postulated that it would reach long-term circulation within the next five years [6]. Experience in the last 2 years has revealed that resurgence as “waves” is common [4], and the overall pattern of the coronavirus pandemic has been a series of increased COVID-19 waves: which gradually declines. Several outbreaks of illnesses have occurred in various parts of the world. Europe entered its third phase in March 2021 [7], and the United States is likely to experience the 4th wave [8]. The reason for the seasonal variation in transmission, duration of immunity, degree of cross-immunity between SARS-CoV-2 and other coronaviruses, intensity and timing of control measures [7], and not the least viral mutation can all explain the wave and gradual decline of cases (waves of the disease). [10]. SARS-CoV-2 has mutated at a pace of about 1–2 mutations per month throughout the current global crisis [11]. A variant of SARS-CoV-2 with the *D614G* mutation emerged in China in late January and early February 2020 and immediately became the dominant form of the virus circulating globally [10]. Subsequently, several mutations were recognized as mutations of concern, naming the UK variant known as 501Y. V1, VOC 202012/01, and B.1.1.7, a South African variant known as the 501Y. V2, or B.1.351, a Brazilian variant known as 501Y. V3 or lineage [11, 12, 13], and the Indian variant B.1.617 (double mutant) [14]. The Indian variant gave rise to India’s second wave and stressed its health system. In terms of test positivity rates (TPRs) and case fatality rates (CFRs), the epidemiologic characteristics seem to differ from those of the first wave [15]. Bangladesh reported the peak of the second wave in late February 2021 and the third wave in June 2021 [16]. In the second wave, death rate increased among the young [17]. The African variant was responsible for more than 80% of the second wave, while the delta variant was responsible for 78% of the third wave [17,18]. Preliminary reports of various SARS-CoV-2 mutant strains show transmissibility, severity, and case mortality diversity. The initial study about the African variant reported no significant correlation between severe disease and outcomes. However, higher transmissibility is possible. The mutant virus behavior in Bangladeshi populations remains largely unknown.

Therefore, this study aimed to systematically and scientifically observe the difference in the presentation and outcome of patients with COVID-19 between the first, second, and third waves.

Additionally, we examined the positivity rate, hospital admission rate, and death rate in outpatient department patients.

## Materials and Methods

This study aimed to examine differences in the epidemiology of COVID-19 in the first, second, and third waves. We aimed to find if there were any variations in the symptoms, disease severity, case fatality, length of hospital stay, frequency of patients requiring oxygen therapy, and referrals to the intensive care unit (ICU). We reviewed the outpatient data for clinic attendance, hospital admission rate, and death in the outpatient department. We examined the virology laboratory data to observe differences in the positivity rate, number of tests, and age and sex. We reviewed inpatient hospital data for rates of hospital admission and death. We found differences in complications of COVID-19 in patients followed up for at least 6 months after hospital discharge during the first, second, and third waves of COVID-19.

### Study area and period

The study was conducted at Dhaka Medical College Hospital between January 01, 2021, and June 20, 2022.

### Study design

We conducted a cross-sectional study and a review of the hospital records. We reviewed the hospital records of outpatients, inpatients, and virology departments. We examined the patients’ medical. Some patients were followed up for at least 6 months after discharge for post-COVID-19 complications. The telephone interviews followed pre-specified telephone interview guidelines.

### Source population

The Dhaka Medical College Hospital inpatient were the source population.

### Study population

The study population included inpatients and patients who called back at least 6 months after follow-up.

### Eligibility criteria

The study included patients’ records, containing important demographics such as age, sex, area of residence, telephone number, complete treatment sheet, admission and discharge date, and at least a brief history. Patients more than 18 years of age, with RT-PCR positive test results irrespective of the severity of the disease admitted between April 2020 and September 2022 were included in this study for analysis and follow-up.

### Sample size determination

The sample size was calculated using the formula below, and the hypothesis was that there was no variation in presentation between the COVID-19 waves in Bangladesh during the pandemic:

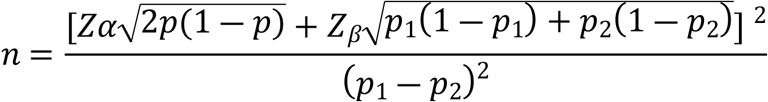

Here, Zα=1.96 Z value of standard normal distribution at 95% confidence interval, P1=prevalence of severe infection in the first wave, P2=Prevalence of severe disease in the second wave, RR=1.5[risk ratio], P2= 0.12; in one study in Bangladesh, the prevalence of severe infection was 11.5% [19], P1=RR×p2=0.18, Zβ=0.84 at 80% power, P= (p1+p2) ÷2=0.15. Thus, the estimated sample size was 240 for each wave.

### Sampling technique and procedure

Cluster sampling methods were used. We compiled a list of all wards where patients with COVID-19 had been admitted. We selected one male ward and one female ward. We subsequently reviewed all records that met the inclusion and exclusion criteria. We selected post-COVID follow-up with non-random sampling methods.

### Study variables

The variables included in the study were

**Demographic variable:** Age of the respondents in years and sex

**Presenting features of COVID-19:** Fever, cough, respiratory distress, sore throat, nausea, vomiting, diarrhea, and body ache.

**Investigations in COVID-19 state:** Total blood count, CRP, D-dimer, creatinine, and ferritin levels.

**COVID-19-severity:** COVID-19 positivity, symptoms in case of COVID-19 positivity (asymptomatic, mild, moderate, severe, or critical)

**Duration:** Duration between symptom onset and hospital admission, length of hospital stay.

**Post-COVID-19 conditions:** Fatigue, cough, respiratory distress, insomnia, etc.

**Outcome-related questions:** Discharged and death. Oxygen requirement, ICU referral **Functional impact:** We examined functional implications in the post-covid state for at least 6 months for overall functional status, fatigue score, and depression score.

**Operational definition:** Confirmed Covid-19, and its different severities were defined according to the WHO and National guidelines [19, 20].

**Case fatality rate:**

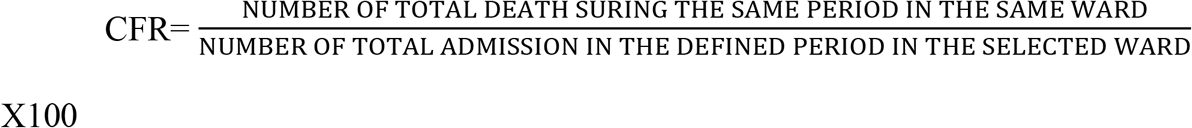

**Duration of hospital stay**: The total period from the day of admission to the day of discharge.

**Waves of infection:**The first, second, and third waves extend between April 2020 to January 2020, February 2021 and May 2021, and June 2021 and September 2021, respectively [21,22].

**Post-COVID-19 condition: According to the Center for Disease Control, USA [23]**

Post-COVID conditions include a wide range of new, returning, or ongoing health problems that people experience after contracting the SARS-CoV-2 causing COVID-19.

**Post-COVID fatigue**: The diagnostic criteria for post-COVID-19 fatigue were developed according to the United States Institute of Medicine symptom criteria [24].

The Chalder Fatigue Scale [25], the Karnofsky Performance Status Scale [26], and the depression scale were used to assess fatigue severity, overall functional status, and depression [27].

### Data collection instrument

For patients admitted to the hospital, data were collected on a data collection sheet. A telephone directory was used for Post COVID-19 follow-up. We collected data from reports of outpatient, inpatient, and virology departments.

### Data Collection procedure

We collected all patients’ files from the hospital record unit in the selected ward. Each file was evaluated by one data collector and researcher. The completed data collecting sheet contains a list of all available information. We obtained no specific procedure for missing data and excluded files that did not contain essential information, such as patient demographics, admission date, and discharge date. We collected the phone number from the hospital records and interviewed the patients for post-covid-19 complications at least 6 months after recovery. We collected data from the outpatient, inpatient, and virology departments using the Excel datasheet, which contains the patient’s name, age, sex, date of attendance, outcome, admission, death, discharge, positive or negative status, etc.

### Data Quality Control

Each data sheet was reviewed by two researchers; disagreements were resolved through discussion. We removed responses where respondents provided contradictory answers. Because each patient was coded and had a specified hospital record number, there was no scope for double responses.

### Data Processing and Analysis

R (v4.1.1) was used to process data. We performed a network analysis with a tidy verse and q graph. Qualitative data with normal distribution were expressed as means (SD), while non-normal data were expressed as medians (IQR). We divided the respondents into groups of first, second, and third waves. We used the chi-square test to calculate quantitative data and one-way ANOVA to calculate quantitative data. A needed we used a post hoc analysis with Bonferroni adjustment, Dunn test, and Tukey’s test. We determined epidemiological trends in the EXECL sheet. The p-value for statistical significance was set at <0.05. We did not impute any missing values, and they were included in the analysis.

### Ethical consideration

The Institutional Ethical Committee of Dhaka Medical College approved this study (ERC-DMC/ECC/2021/55). As we mainly reviewed the hospital data, there was no need for written consent. The patients provided verbal consent for post-covid follow-up.

## Results

We reviewed 1766 patient records and included 1597 patients’ data for analysis in this mixed study of retrospective hospital review of hospital data and cross-sectional follow-up of patients admitted for post-COVID-19 complications. From these patients we followed 600 patients over the telephone for post-COVID-19 complications. Between April 2020 and September 2021, and 38578 patients visited the outpatient department, 24501 patients were admitted, 57857 were tested for treatment at the virology department (Suppliment1).

The mean age (SD) of the hospital-admitted patients was 47.91(15.67) years, with patients in the and third waves being higher than those in the first wave (p <0.001). Patients above 60 years were mainly involved in the first wave 144 (54.8%), the 40–60 age group was primarily involved in the second wave 139 (39.8%), and those above 60 years were more likely to be infected in the third wave 161 (43.3%).

Men were predominantly admitted 968 (60.68%). However, in the second wave, the number of women, 191 (54.7%), was higher.

Fever was present in 1366 (85.6%) patients and did not differ among the three waves (p = 0.1). Cough was more prevalent in the second 288 (65.5%) and third waves 290 (78%) than in the first wave 522 (59.7%). Running nose was most prevalent in the first wave, 143 (16.4%), and least prevalent in the second wave, 16 (4.6%). Respiratory distress was the most prevalent in the third wave, 251 (67.1%), and the least prevalent in the first wave, 403 (46.1%). Sore throat occurred more prevalent in the first wave 198 (22.7%) than in the other waves. Diarrhea 60 (16.1) and vomiting 59 (15.9%) were the most prevalent in the third wave. Anosmia was more prevalent in the third wave 116 (31.2%) than in other waves and less frequently in the second wave 27 (7.8%) than in other waves. However, headache was more prevalent in the third 82 (22.5%) and first wave 145 (16.6%) than in the second wave. Body aches occurred more commonly in the second 87 (25%) and third 92 (24.6%) waves than in the first wave. (Table 1)

**Table 1:**
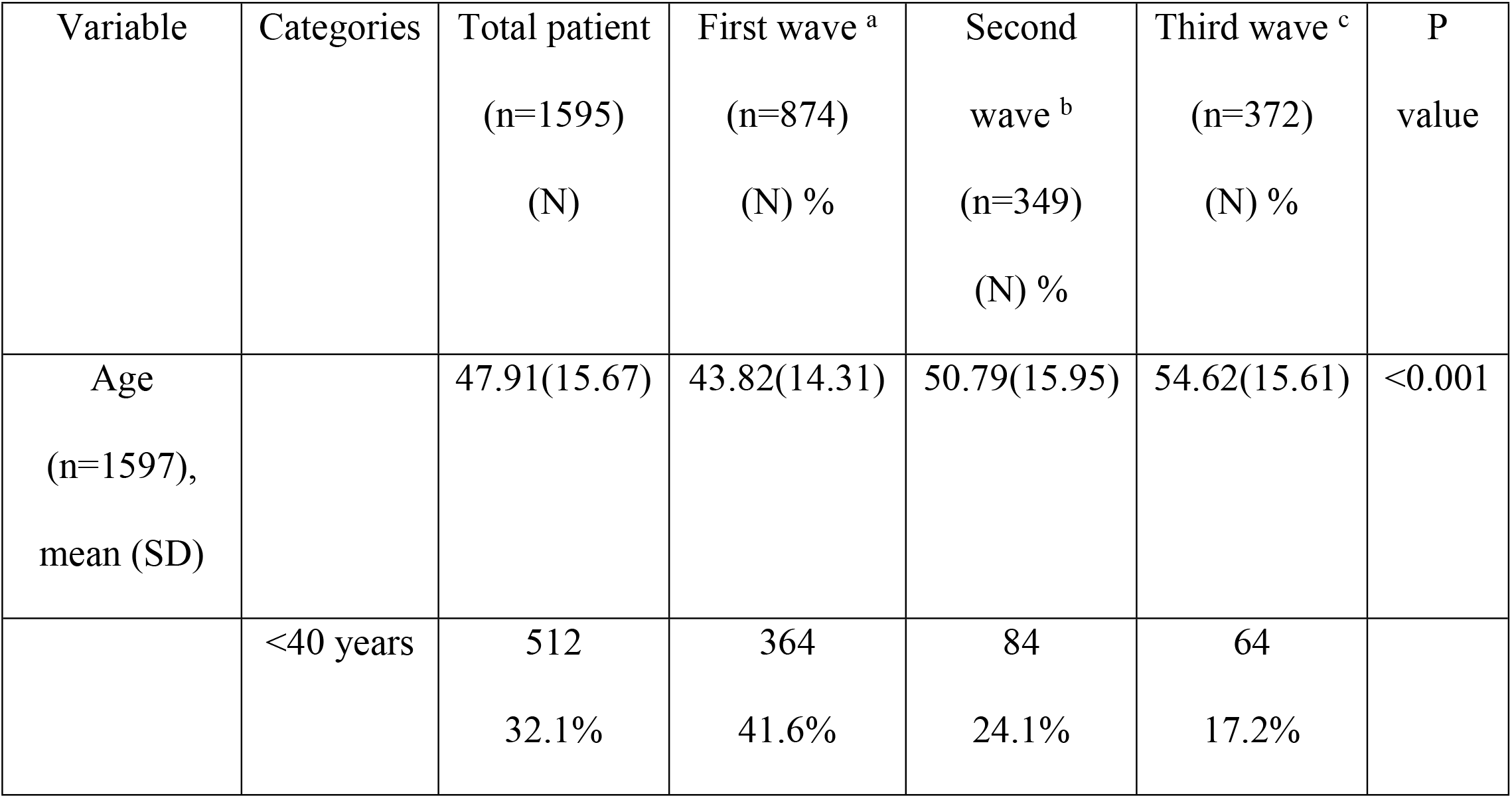
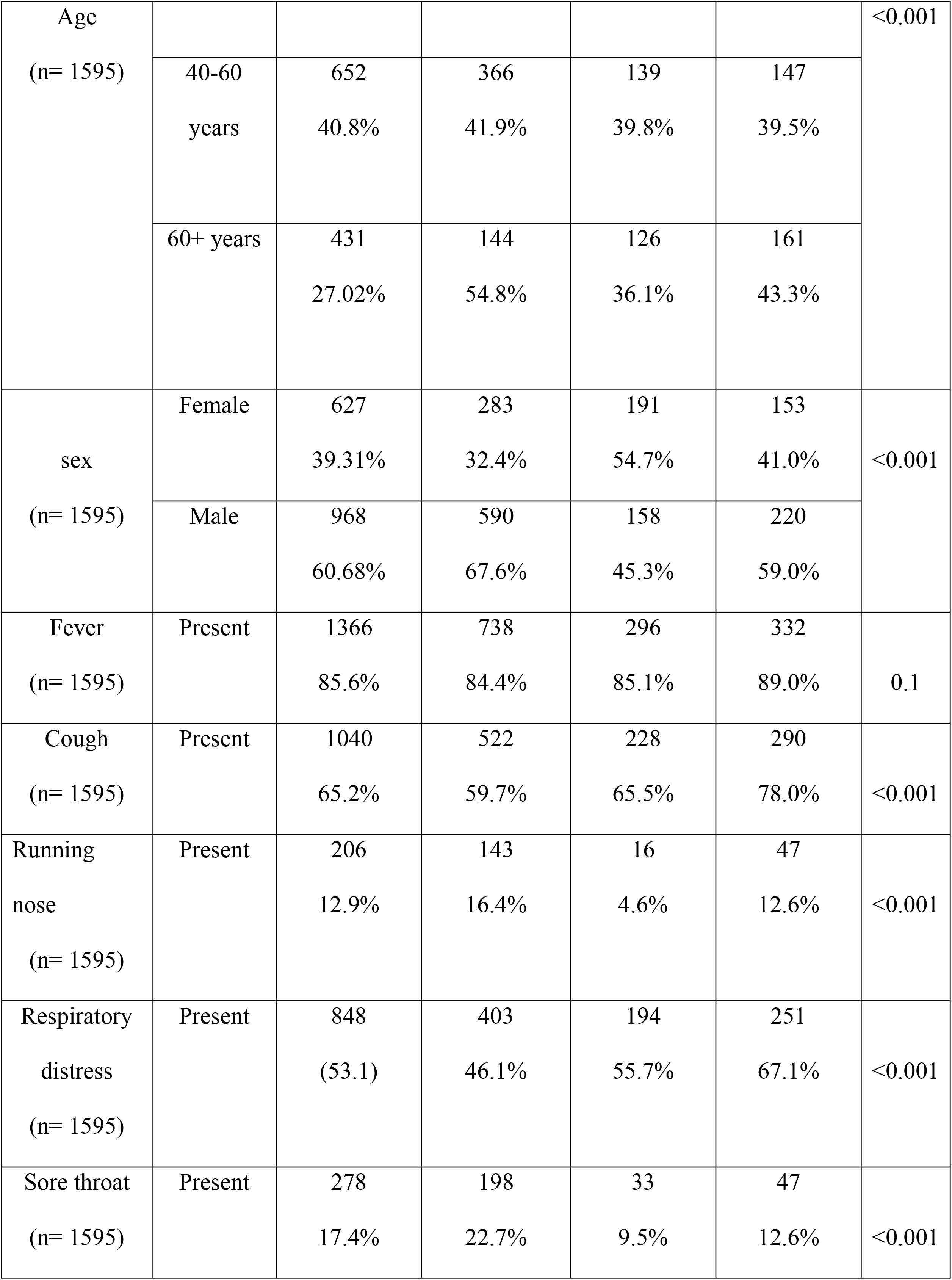
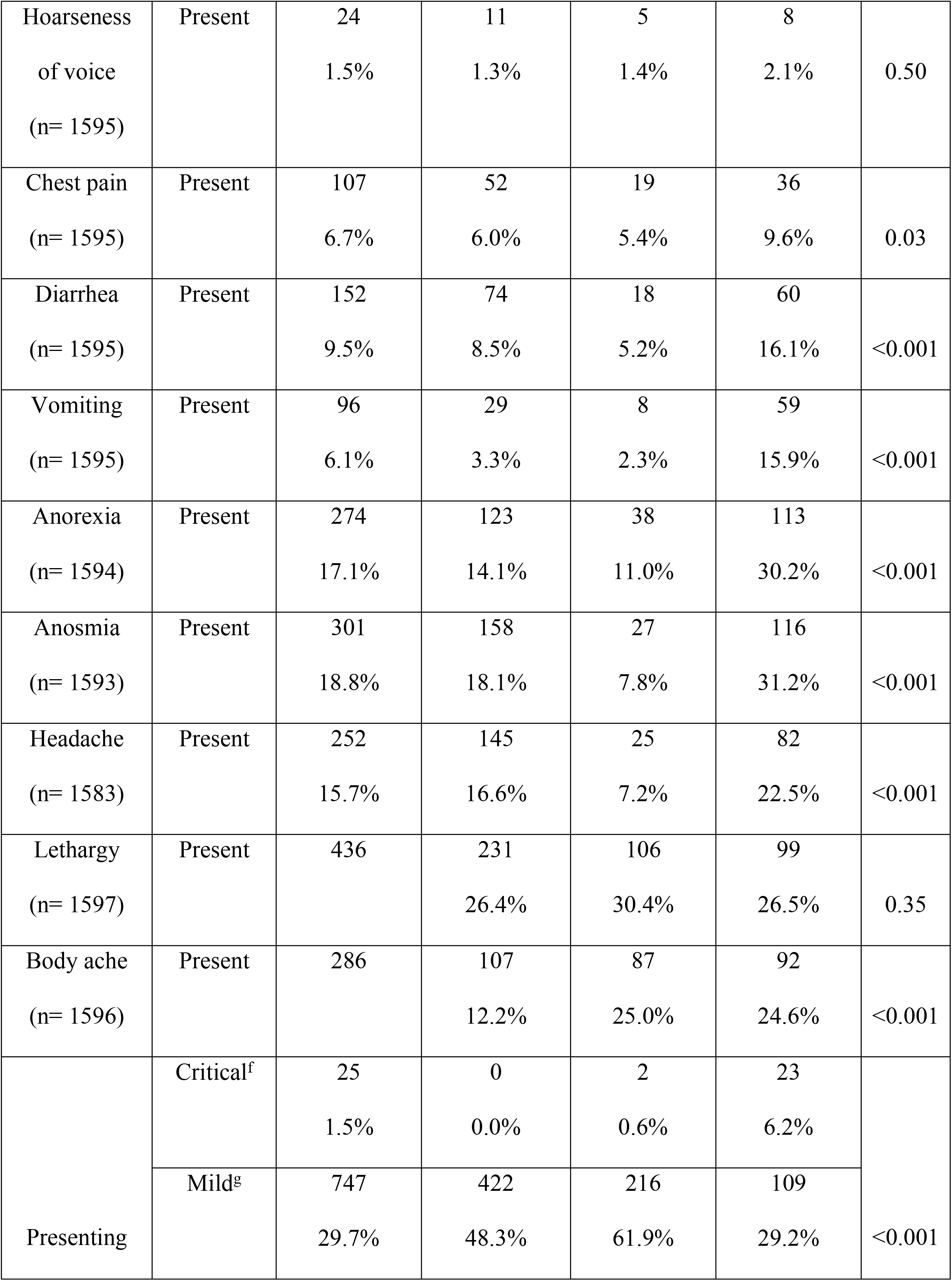
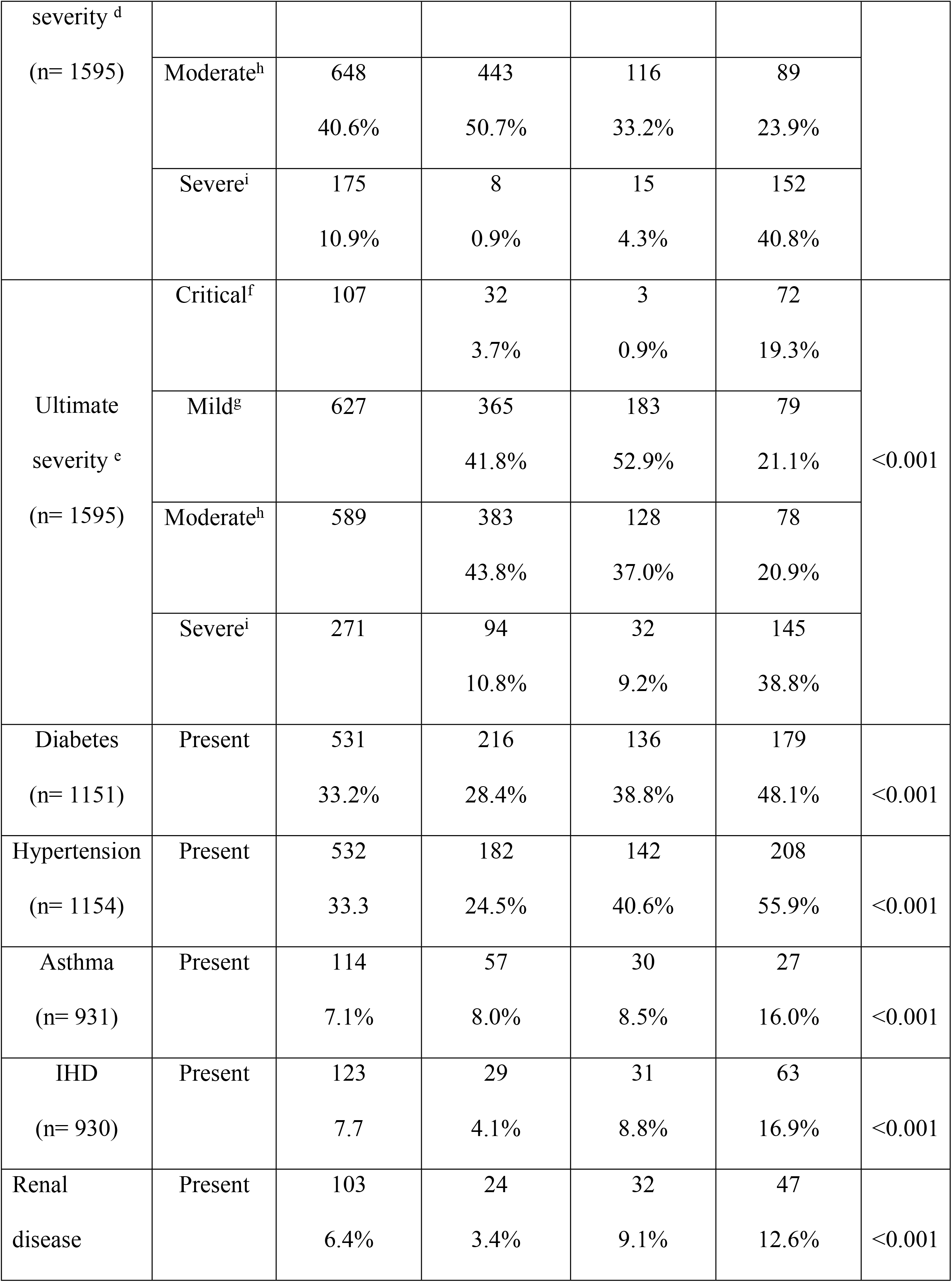
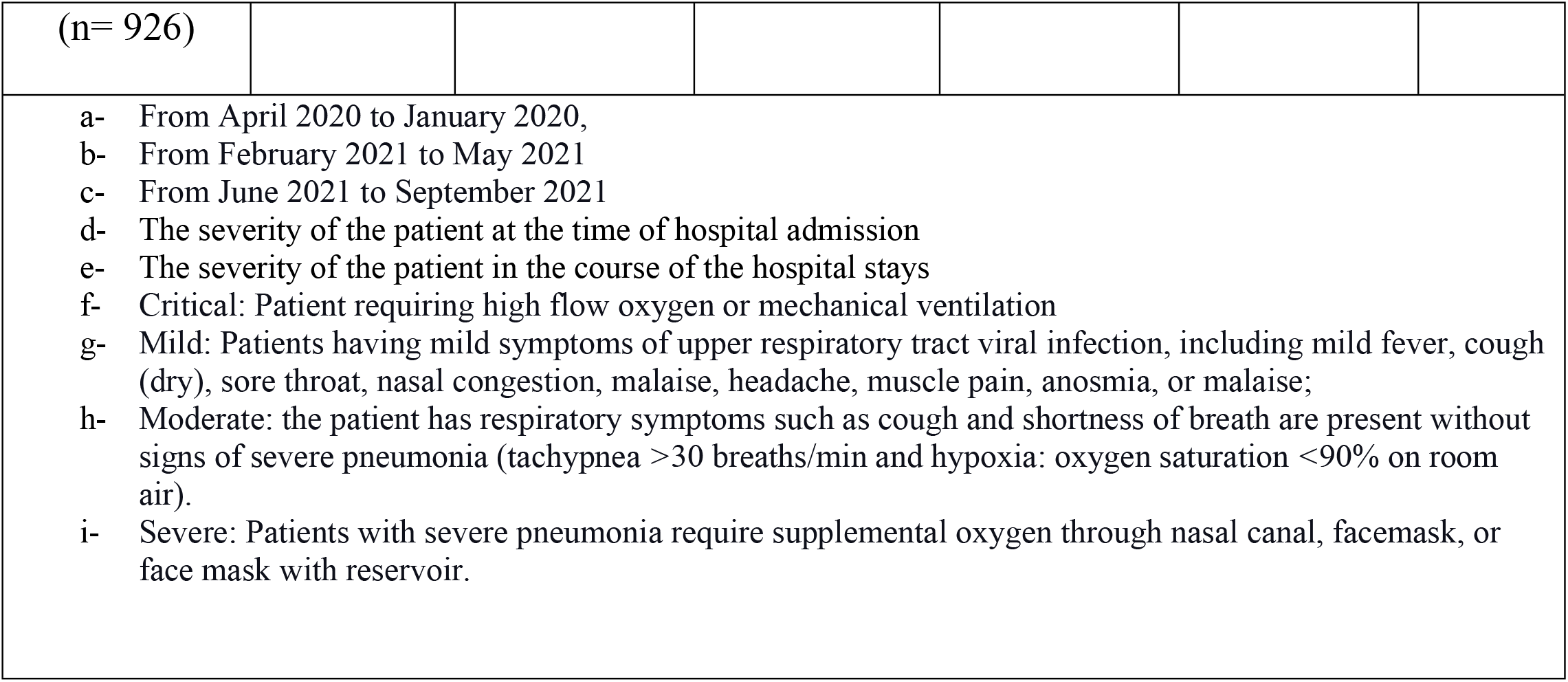
Demographic characteristics and co-morbidity of the admitted patients in the three waves.

| Variable | Categories | Total patient<br>(n=1595)<br>(N) | First wave <sup>a</sup><br>(n=874)<br>(N) % | Second<br>wave <sup>b</sup><br>(n=349)<br>(N) % | Third wave <sup>c</sup><br>(n=372)<br>(N) % | P<br>value |
| --- | --- | --- | --- | --- | --- | --- |
| Age<br>(n=1597),<br>mean (SD) |  | 47.91(15.67) | 43.82(14.31) | 50.79(15.95) | 54.62(15.61) | <0.001 |
|  | <40 years | 512<br>32.1% | 364<br>41.6% | 84<br>24.1% | 64<br>17.2% |  |
| Age<br>(n= 1595) |  |  |  |  |  | <0.001 |
|  | 40-60<br>years | 652<br>40.8% | 366<br>41.9% | 139<br>39.8% | 147<br>39.5% |  |
|  | 60+ years | 431<br>27.02% | 144<br>54.8% | 126<br>36.1% | 161<br>43.3% |  |
| sex<br>(n= 1595) | Female | 627<br>39.31% | 283<br>32.4% | 191<br>54.7% | 153<br>41.0% | <0.001 |
|  | Male | 968<br>60.68% | 590<br>67.6% | 158<br>45.3% | 220<br>59.0% |  |
| Fever<br>(n= 1595) | Present | 1366<br>85.6% | 738<br>84.4% | 296<br>85.1% | 332<br>89.0% | 0.1 |
| Cough<br>(n= 1595) | Present | 1040<br>65.2% | 522<br>59.7% | 228<br>65.5% | 290<br>78.0% | <0.001 |
| Running<br>nose<br>(n= 1595) | Present | 206<br>12.9% | 143<br>16.4% | 16<br>4.6% | 47<br>12.6% | <0.001 |
| Respiratory<br>distress<br>(n= 1595) | Present | 848<br>(53.1) | 403<br>46.1% | 194<br>55.7% | 251<br>67.1% | <0.001 |
| Sore throat<br>(n= 1595) | Present | 278<br>17.4% | 198<br>22.7% | 33<br>9.5% | 47<br>12.6% | <0.001 |
| Hoarseness<br>of voice<br>(n= 1595) | Present | 24<br>1.5% | 11<br>1.3% | 5<br>1.4% | 8<br>2.1% | 0.50 |
| Chest pain<br>(n= 1595) | Present | 107<br>6.7% | 52<br>6.0% | 19<br>5.4% | 36<br>9.6% | 0.03 |
| Diarrhea<br>(n= 1595) | Present | 152<br>9.5% | 74<br>8.5% | 18<br>5.2% | 60<br>16.1% | <0.001 |
| Vomiting<br>(n= 1595) | Present | 96<br>6.1% | 29<br>3.3% | 8<br>2.3% | 59<br>15.9% | <0.001 |
| Anorexia<br>(n= 1594) | Present | 274<br>17.1% | 123<br>14.1% | 38<br>11.0% | 113<br>30.2% | <0.001 |
| Anosmia<br>(n= 1593) | Present | 301<br>18.8% | 158<br>18.1% | 27<br>7.8% | 116<br>31.2% | <0.001 |
| Headache<br>(n= 1583) | Present | 252<br>15.7% | 145<br>16.6% | 25<br>7.2% | 82<br>22.5% | <0.001 |
| Lethargy<br>(n= 1597) | Present | 436 | 231<br>26.4% | 106<br>30.4% | 99<br>26.5% | 0.35 |
| Body ache<br>(n= 1596) | Present | 286 | 107<br>12.2% | 87<br>25.0% | 92<br>24.6% | <0.001 |
| Presenting | Critical<sup>f</sup> | 25<br>1.5% | 0<br>0.0% | 2<br>0.6% | 23<br>6.2% | <0.001 |
|  | Mild<sup>g</sup> | 747<br>29.7% | 422<br>48.3% | 216<br>61.9% | 109<br>29.2% |  |
| severity <sup>d</sup><br>(n= 1595) |  |  |  |  |  |  |
|  | Moderate<sup>h</sup> | 648<br>40.6% | 443<br>50.7% | 116<br>33.2% | 89<br>23.9% |  |
|  | Severe<sup>i</sup> | 175<br>10.9% | 8<br>0.9% | 15<br>4.3% | 152<br>40.8% |  |
| Ultimate<br>severity <sup>e</sup><br>(n= 1595) | Critical<sup>f</sup> | 107 | 32<br>3.7% | 3<br>0.9% | 72<br>19.3% | <0.001 |
|  | Mild<sup>g</sup> | 627 | 365<br>41.8% | 183<br>52.9% | 79<br>21.1% |  |
|  | Moderate<sup>h</sup> | 589 | 383<br>43.8% | 128<br>37.0% | 78<br>20.9% |  |
|  | Severe<sup>i</sup> | 271 | 94<br>10.8% | 32<br>9.2% | 145<br>38.8% |  |
| Diabetes<br>(n= 1151) | Present | 531<br>33.2% | 216<br>28.4% | 136<br>38.8% | 179<br>48.1% | <0.001 |
| Hypertension<br>(n= 1154) | Present | 532<br>33.3 | 182<br>24.5% | 142<br>40.6% | 208<br>55.9% | <0.001 |
| Asthma<br>(n= 931) | Present | 114<br>7.1% | 57<br>8.0% | 30<br>8.5% | 27<br>16.0% | <0.001 |
| IHD<br>(n= 930) | Present | 123<br>7.7 | 29<br>4.1% | 31<br>8.8% | 63<br>16.9% | <0.001 |
| Renal<br>disease | Present | 103<br>6.4% | 24<br>3.4% | 32<br>9.1% | 47<br>12.6% | <0.001 |
| (n= 926) |  |  |  |  |  |  |
| a- From April 2020 to January 2020,<br>b- From February 2021 to May 2021<br>c- From June 2021 to September 2021<br>d- The severity of the patient at the time of hospital admission<br>e- The severity of the patient in the course of the hospital stays<br>f- Critical: Patient requiring high flow oxygen or mechanical ventilation<br>g- Mild: Patients having mild symptoms of upper respiratory tract viral infection, including mild fever, cough (dry), sore throat, nasal congestion, malaise, headache, muscle pain, anosmia, or malaise;<br>h- Moderate: the patient has respiratory symptoms such as cough and shortness of breath are present without signs of severe pneumonia (tachypnea >30 breaths/min and hypoxia: oxygen saturation <90% on room air).<br>i- Severe: Patients with severe pneumonia require supplemental oxygen through nasal canal, facemask, or face mask with reservoir. |  |  |  |  |  |  |

Patients that presented in a critical condition 23 (6.2%) and severe disease 152 (40.8%) were more prevalent in the third wave than in other waves. In the second wave, most cases were mild 216 (61.9%). During the hospital stay, the severity of the patient’s severity changed, and in the third wave, 72 (19.3%) critical and 145 (38.8%) severe diseases emerged. Mild-to-moderate illness was more common in the first and second waves than in the third wave.

Diabetes was more common among admitted patients in the second 136 (38.8%) and third waves 179 (48.1%) than in the first wave. Hypertension was prevalent in the second 142 (40.6%) and third wave 208 (55.9%). Asthma 27 (16%), ischemic heart disease 63 (16.9%), and renal disease 47 (12.6%) were the most prevalent in the third wave (Table 1).

### Symptoms Cluster in COVID-19

We found three clusters of symptoms among the admitted COVID patients: 1. Fever, cough, and respiratory distress. 2. Anosmia, headache, and anorexia 3. Sore throat and running nose. The symptom clusters differed for the three waves. In the first wave, we found three clusters; 1.

Respiratory distress and cough. 2. Anosmia and anorexia. 3. Running nose and sore throat. In the second wave, we found a cluster of headaches, running nose, sore throat, and hoarseness of voice. In the third wave, we found three clusters: 1. anorexia and anosmia, 2. fever and cough, and 3. headache, diarrhea, and vomiting. (Figure 1)

**Figure 1:**
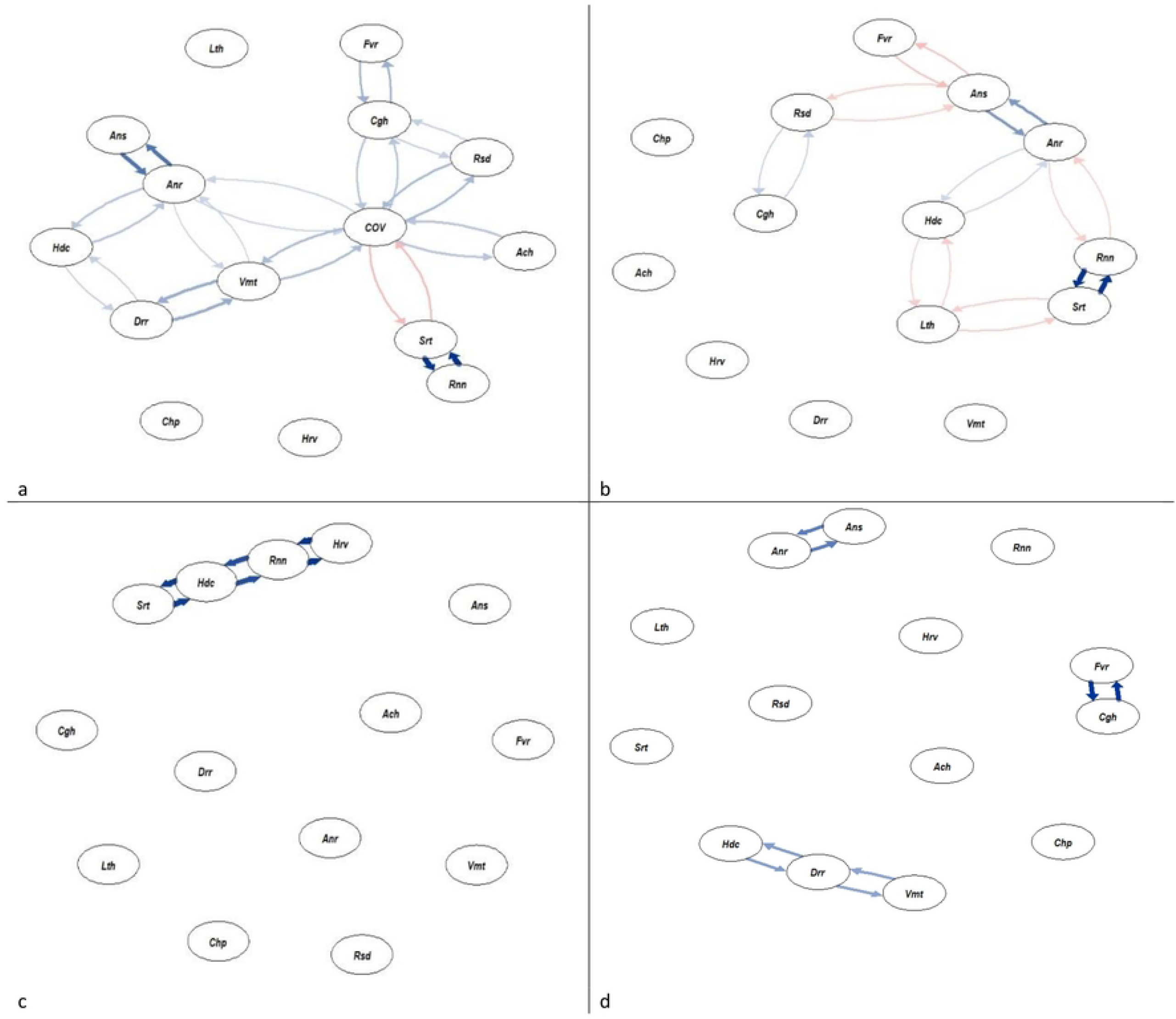
Symptoms clusters in COVID-19. a. According to general clusters of symptoms, We found three clusters of symptoms among the admitted patients with COVID-19: I. Fever, cough, and respiratory distress. 2. Anosmia, headache, and anorexia, 3. Sore throat and running nose. b. In the first wave, we found three clusters I. Respiratory distress and cough, 2. Anosmia anorexia, and 3. Running nose and sore throat. c. In the second wave, we found a cluster headache, running nose, sore throat, and hoarseness of voice. d. In the third wave, we found three clusters I. Anorexia and anosmia, 2. Fever and cough, and 3. headache, diarrhea, and vomiting.We did the network analysis with tidy verse and q graph using R (v4.1.l).(cov-COVID-19, Fvr-Fever, Cgh-cough, Ron-Running nose, Rsd-Respiratory distress, Srt-Sore throat, Hrv-Hoarseness of voice, Chp-Chest pain, Orr-Diarrhea, Vmt-vomiting, Anr-Anorexia, Ans: Anosmia, Hdc-Headache, Lth-Lethargy, Ach-Body ache)

The white blood cell count was high in the second wave, 18.65(14.5–24), and low in the third wave. The platelet count was low in the third wave 78 (68–86). CRP was much higher in the third wave 39 (13–85) and D-dimer 1.52 (0.63–3.56), and ferritin was high in the second wave 1468 (973–2050). (Table 2)

**Table 2:**
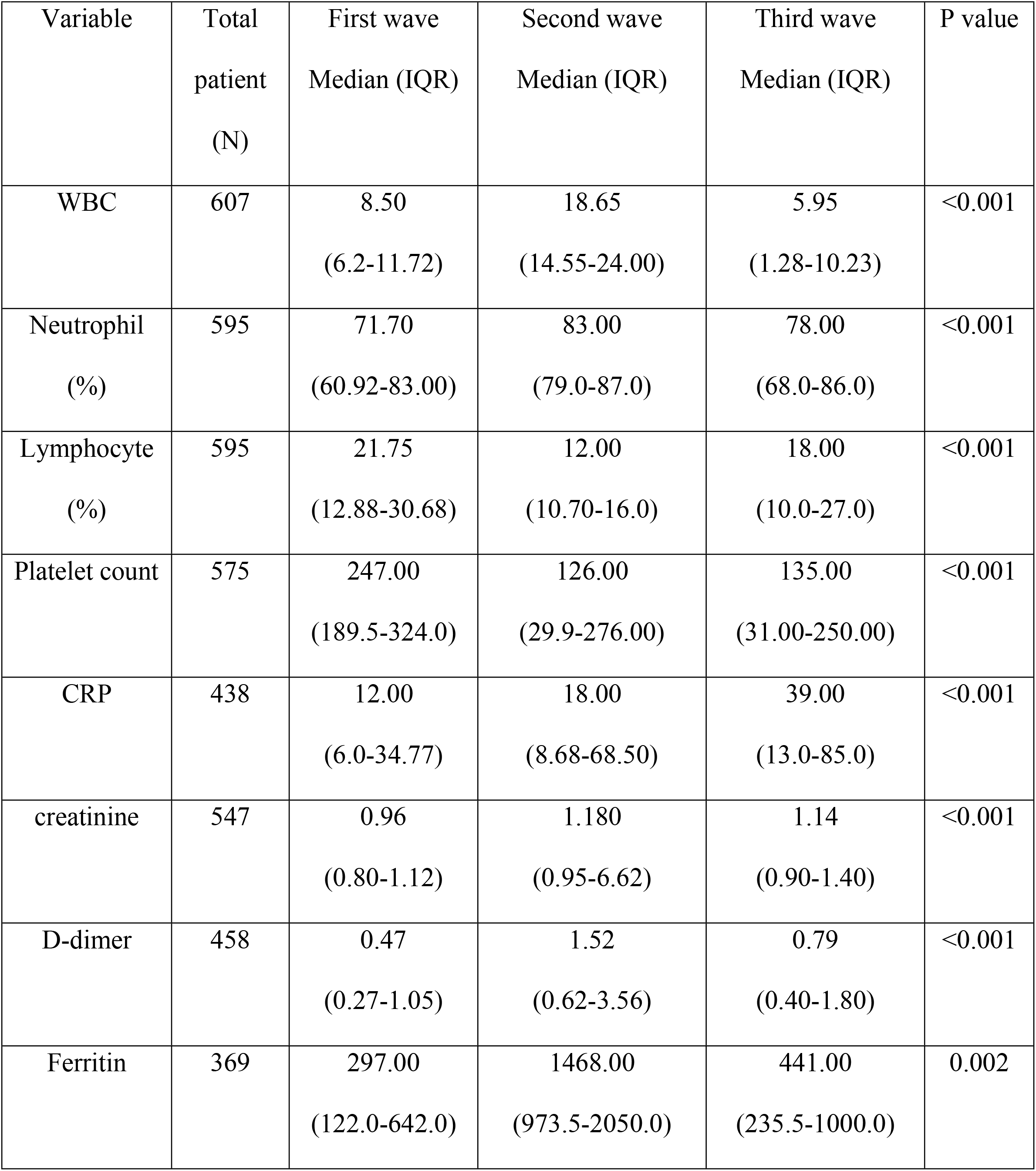
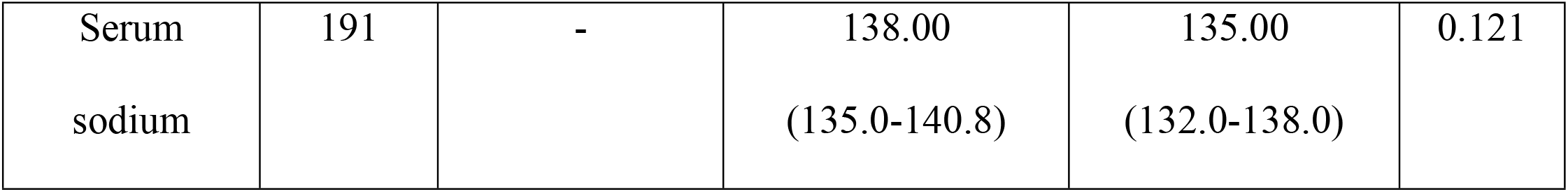
Investigation profile of the admitted patients in the three waves.

The patient developed a more severe disease in the third wave, and the duration of hospital stay was longer in the first wave 12 (8–20). (Table 3)

**Table 3:** Outcome of the admitted patient in three waves.

| Variable | Categories | Total patient (N) | First wave (N)<br>% | Second wave (N)<br>% | Third wave (N)<br>% | P value |
| --- | --- | --- | --- | --- | --- | --- |
| Ultimate severity | Critical | 107 | 32<br>29.9% | 3<br>2.8% | 72<br>67.3% | <0.001 |
|  | Mild | 627 | 365<br>58.2% | 183<br>29.2% | 79<br>12.6% |  |
|  | Moderate | 589 | 383<br>65.0% | 128<br>21.7% | 78<br>13.2% |  |
|  | Severe | 271 | 94<br>34.7% | 32<br>11.8% | 145<br>53.5% |  |
| Duration of hospital stay |  |  | 13.0<br>(8.0- | 12.0<br>(8.0-12.0) | 7.0<br>(4.0-11.0) | <0.001 |
|  |  |  | 20.0) |  |  |  |
| Interval<br>symptom<br>onset and<br>admission |  |  | 5.0<br>(4.0-6.0) | 5.0<br>(5.0-6.0) | 5.0<br>(4.0-6.0) | <0.001 |

Confirmed COVID-19 cases were higher (15.7%) in the second wave than in the other waves, the admission rate 73.7%, and the death rate 1.4% were higher in the second wave than in other waves. The proportion of confirmed COVID-19 cases was higher among women in all waves. In the third wave, the admission rate among women 61.1% was higher than among men. The number of positive cases was higher in those below 40 years in the first wave 13.9% and above 60 years in the third wave 18.2%. The admission rate of patients below 40 years was lower in the third wave 33.7%. In the second wave, the admission rate among the 40–60-year age group 79% and above 60 years age group 91.4% was higher than in other waves. In the second wave, the death rate was higher among men 1.4% and women, 1.3% In the second wave, the death rate was high in the 40–60-year age group and above 60-year age group. The admission rate was higher in patients aged above 60 years in all three waves.

52% of patient were admitted in the first wave and 75% received more oxygen in the third wave. Death was more common 51% in the first wave.

The patient was tested more frequently in the first and third waves than in the second wave. The positivity was high in the third wave 22.8%. In the third wave, the positivity rate was high among women 24.3%. The incidence rate of patients above 60 years was high in the first and third waves 27.3%. (Table 4)

**Table 4:**
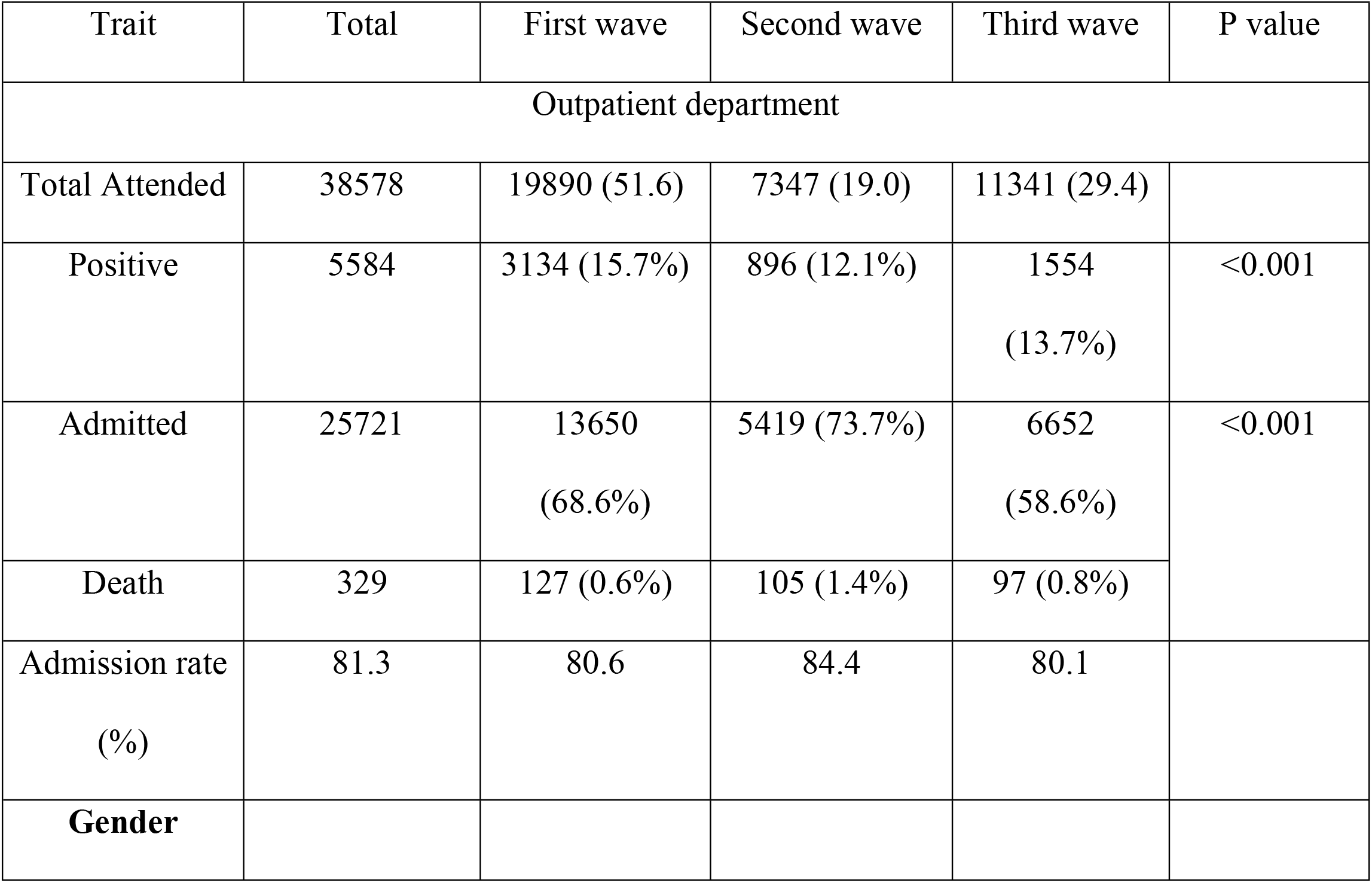
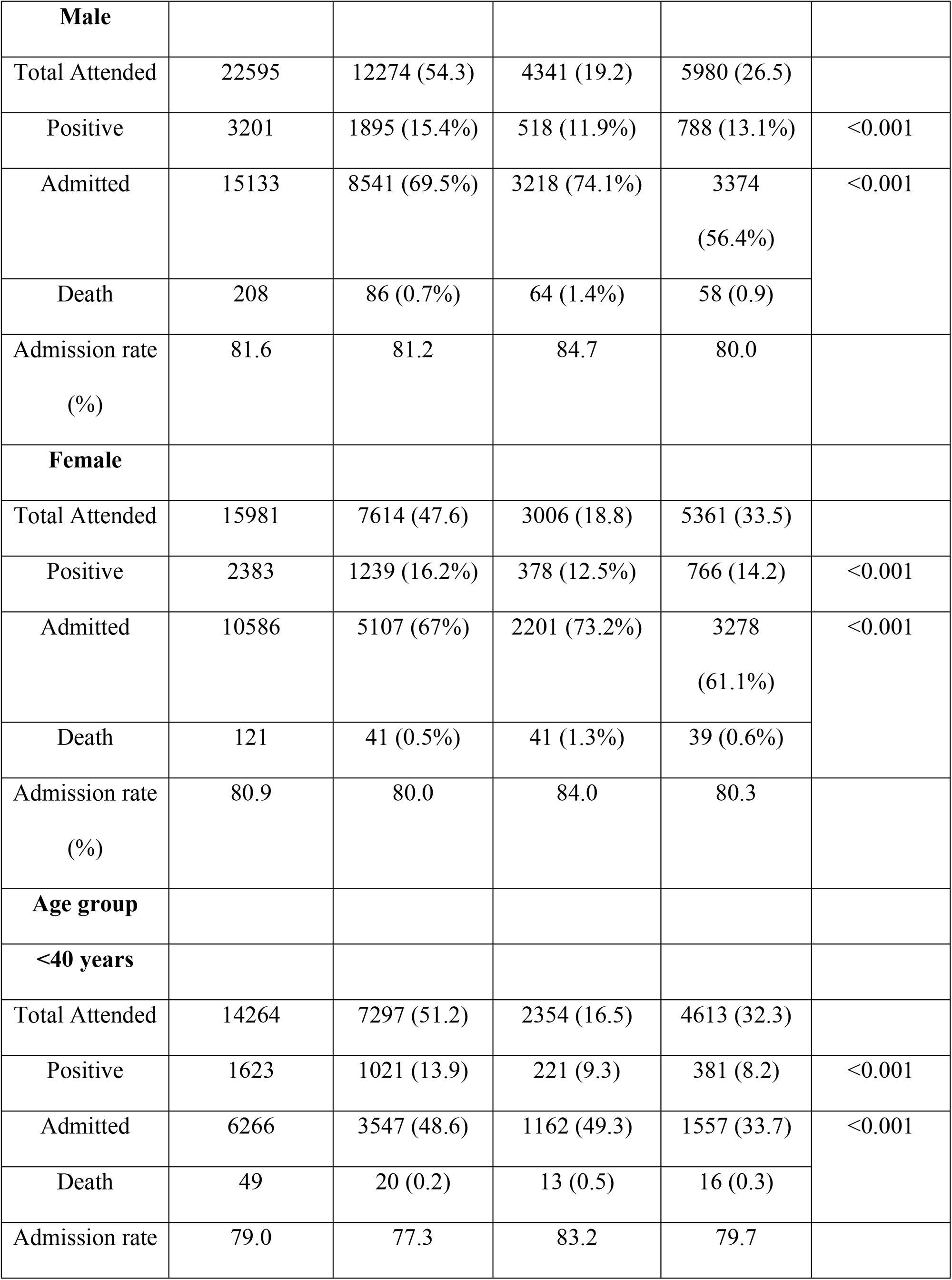
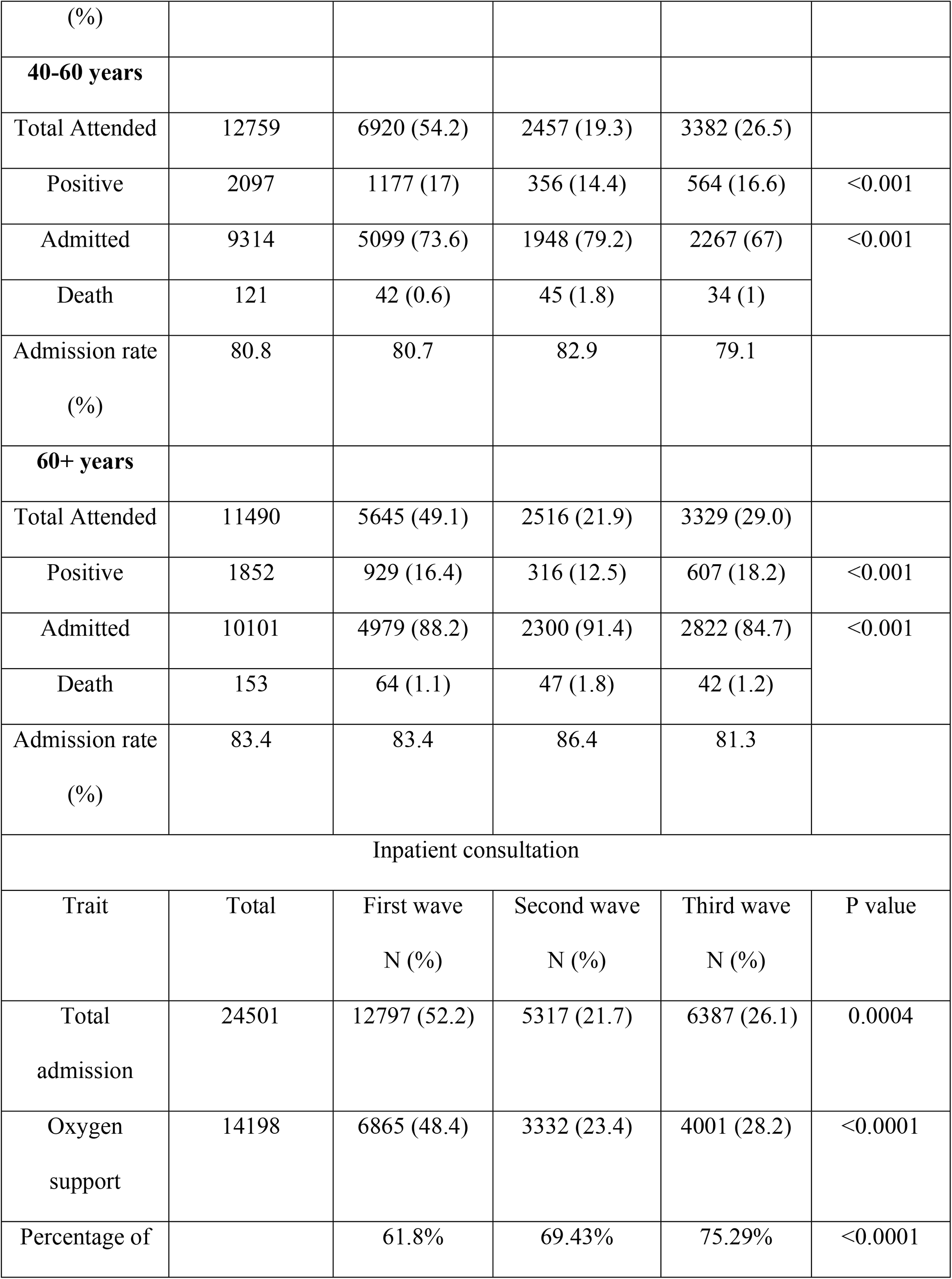
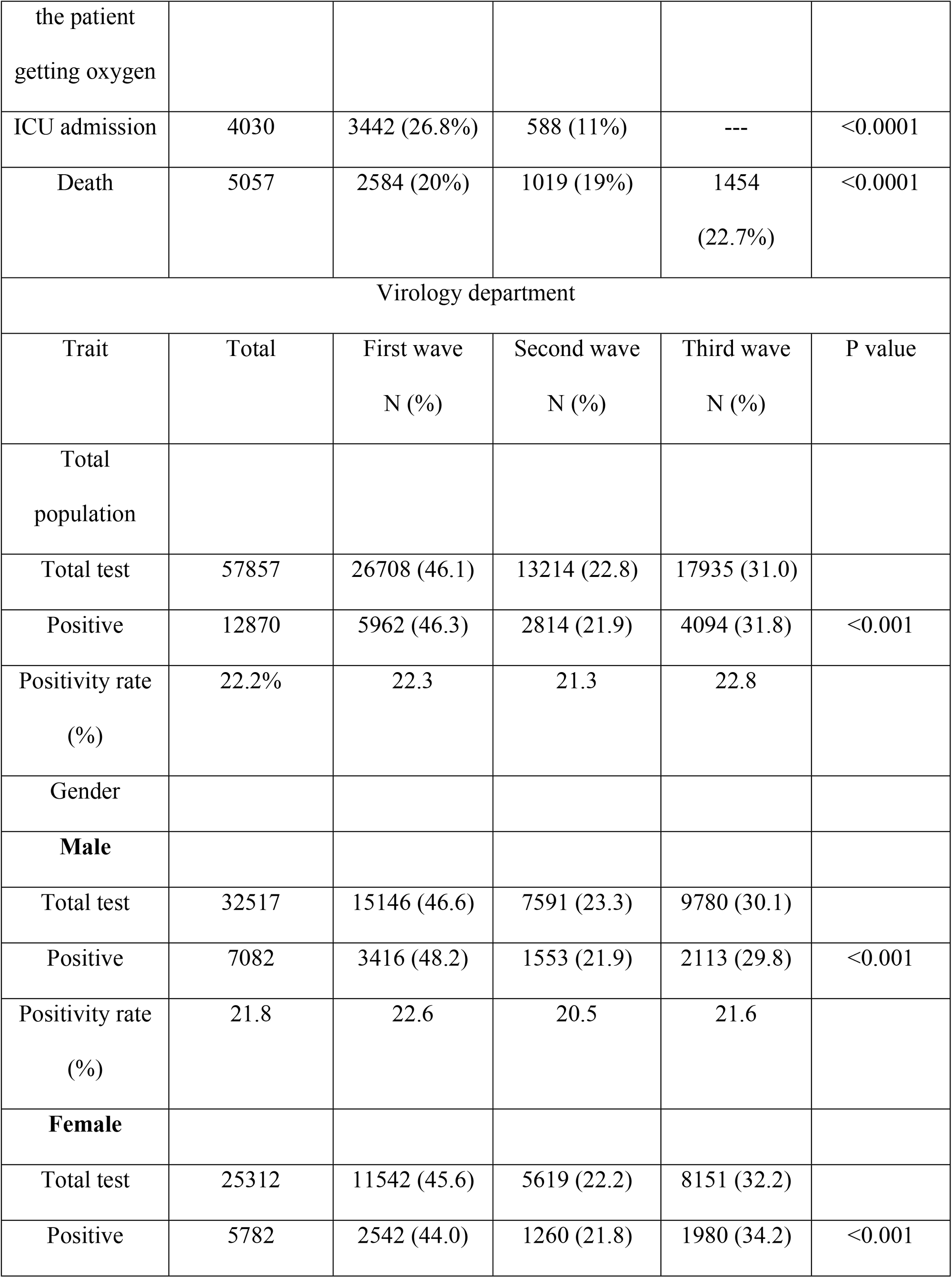
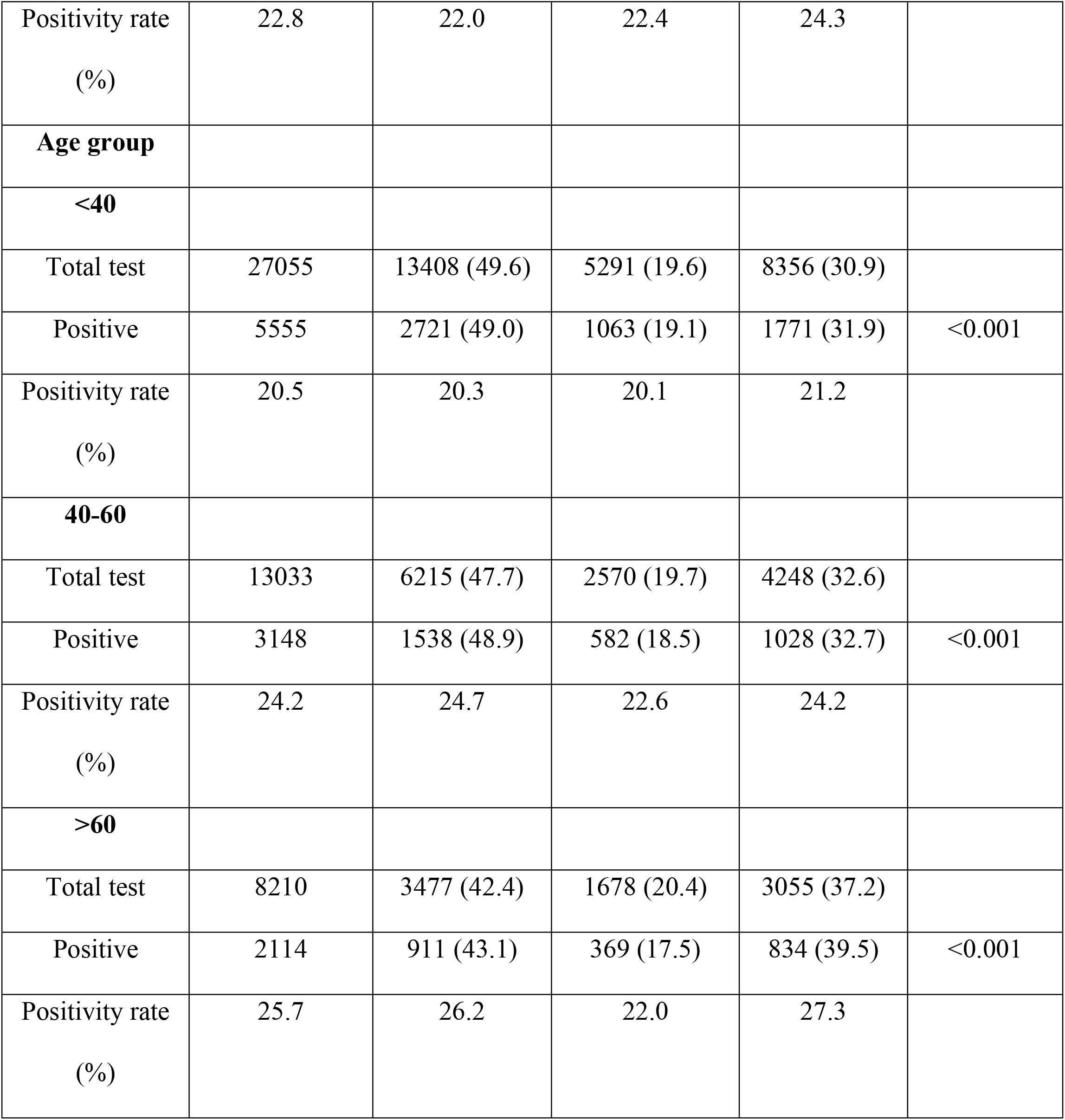
Trend of outpatient, inpatient and virology department consultation in three waves.

| Trait | Total | First wave | Second wave | Third wave | P value |
| --- | --- | --- | --- | --- | --- |
| Outpatient department |  |  |  |  |  |
| Total Attended | 38578 | 19890 (51.6) | 7347 (19.0) | 11341 (29.4) |  |
| Positive | 5584 | 3134 (15.7%) | 896 (12.1%) | 1554<br>(13.7%) | <0.001 |
| Admitted | 25721 | 13650<br>(68.6%) | 5419 (73.7%) | 6652<br>(58.6%) | <0.001 |
| Death | 329 | 127 (0.6%) | 105 (1.4%) | 97 (0.8%) |  |
| Admission rate<br>(%) | 81.3 | 80.6 | 84.4 | 80.1 |  |
| **Gender** |  |  |  |  |  |
| **Male** |  |  |  |  |  |
| Total Attended | 22595 | 12274 (54.3) | 4341 (19.2) | 5980 (26.5) |  |
| Positive | 3201 | 1895 (15.4%) | 518 (11.9%) | 788 (13.1%) | <0.001 |
| Admitted | 15133 | 8541 (69.5%) | 3218 (74.1%) | 3374 (56.4%) | <0.001 |
| Death | 208 | 86 (0.7%) | 64 (1.4%) | 58 (0.9%) |  |
| Admission rate (%) | 81.6 | 81.2 | 84.7 | 80.0 |  |
| **Female** |  |  |  |  |  |
| Total Attended | 15981 | 7614 (47.6) | 3006 (18.8) | 5361 (33.5) |  |
| Positive | 2383 | 1239 (16.2%) | 378 (12.5%) | 766 (14.2) | <0.001 |
| Admitted | 10586 | 5107 (67%) | 2201 (73.2%) | 3278 (61.1%) | <0.001 |
| Death | 121 | 41 (0.5%) | 41 (1.3%) | 39 (0.6%) |  |
| Admission rate (%) | 80.9 | 80.0 | 84.0 | 80.3 |  |
| **Age group** |  |  |  |  |  |
| **<40 years** |  |  |  |  |  |
| Total Attended | 14264 | 7297 (51.2) | 2354 (16.5) | 4613 (32.3) |  |
| Positive | 1623 | 1021 (13.9) | 221 (9.3) | 381 (8.2) | <0.001 |
| Admitted | 6266 | 3547 (48.6) | 1162 (49.3) | 1557 (33.7) | <0.001 |
| Death | 49 | 20 (0.2) | 13 (0.5) | 16 (0.3) |  |
| Admission rate | 79.0 | 77.3 | 83.2 | 79.7 |  |
| (%) |  |  |  |  |  |
| **40-60 years** |  |  |  |  |  |
| Total Attended | 12759 | 6920 (54.2) | 2457 (19.3) | 3382 (26.5) |  |
| Positive | 2097 | 1177 (17) | 356 (14.4) | 564 (16.6) | <0.001 |
| Admitted | 9314 | 5099 (73.6) | 1948 (79.2) | 2267 (67) | <0.001 |
| Death | 121 | 42 (0.6) | 45 (1.8) | 34 (1) |  |
| Admission rate<br>(%) | 80.8 | 80.7 | 82.9 | 79.1 |  |
| **60+ years** |  |  |  |  |  |
| Total Attended | 11490 | 5645 (49.1) | 2516 (21.9) | 3329 (29.0) |  |
| Positive | 1852 | 929 (16.4) | 316 (12.5) | 607 (18.2) | <0.001 |
| Admitted | 10101 | 4979 (88.2) | 2300 (91.4) | 2822 (84.7) | <0.001 |
| Death | 153 | 64 (1.1) | 47 (1.8) | 42 (1.2) |  |
| Admission rate<br>(%) | 83.4 | 83.4 | 86.4 | 81.3 |  |
| **Inpatient consultation** |  |  |  |  |  |
| Trait | Total | First wave<br>N (%) | Second wave<br>N (%) | Third wave<br>N (%) | P value |
| Total admission | 24501 | 12797 (52.2) | 5317 (21.7) | 6387 (26.1) | 0.0004 |
| Oxygen support | 14198 | 6865 (48.4) | 3332 (23.4) | 4001 (28.2) | <0.0001 |
| Percentage of |  | 61.8% | 69.43% | 75.29% | <0.0001 |
| the patient<br>getting oxygen |  |  |  |  |  |
| ICU admission | 4030 | 3442 (26.8%) | 588 (11%) | --- | <0.0001 |
| Death | 5057 | 2584 (20%) | 1019 (19%) | 1454<br>(22.7%) | <0.0001 |
| Virology department |  |  |  |  |  |
| Trait | Total | First wave<br>N (%) | Second wave<br>N (%) | Third wave<br>N (%) | P value |
| Total<br>population |  |  |  |  |  |
| Total test | 57857 | 26708 (46.1) | 13214 (22.8) | 17935 (31.0) |  |
| Positive | 12870 | 5962 (46.3) | 2814 (21.9) | 4094 (31.8) | <0.001 |
| Positivity rate<br>(%) | 22.2% | 22.3 | 21.3 | 22.8 |  |
| Gender |  |  |  |  |  |
| **Male** |  |  |  |  |  |
| Total test | 32517 | 15146 (46.6) | 7591 (23.3) | 9780 (30.1) |  |
| Positive | 7082 | 3416 (48.2) | 1553 (21.9) | 2113 (29.8) | <0.001 |
| Positivity rate<br>(%) | 21.8 | 22.6 | 20.5 | 21.6 |  |
| **Female** |  |  |  |  |  |
| Total test | 25312 | 11542 (45.6) | 5619 (22.2) | 8151 (32.2) |  |
| Positive | 5782 | 2542 (44.0) | 1260 (21.8) | 1980 (34.2) | <0.001 |
| Positivity rate<br>(%) | 22.8 | 22.0 | 22.4 | 24.3 |  |
| **Age group** |  |  |  |  |  |
| **<40** |  |  |  |  |  |
| Total test | 27055 | 13408 (49.6) | 5291 (19.6) | 8356 (30.9) |  |
| Positive | 5555 | 2721 (49.0) | 1063 (19.1) | 1771 (31.9) | <0.001 |
| Positivity rate<br>(%) | 20.5 | 20.3 | 20.1 | 21.2 |  |
| **40-60** |  |  |  |  |  |
| Total test | 13033 | 6215 (47.7) | 2570 (19.7) | 4248 (32.6) |  |
| Positive | 3148 | 1538 (48.9) | 582 (18.5) | 1028 (32.7) | <0.001 |
| Positivity rate<br>(%) | 24.2 | 24.7 | 22.6 | 24.2 |  |
| **>60** |  |  |  |  |  |
| Total test | 8210 | 3477 (42.4) | 1678 (20.4) | 3055 (37.2) |  |
| Positive | 2114 | 911 (43.1) | 369 (17.5) | 834 (39.5) | <0.001 |
| Positivity rate<br>(%) | 25.7 | 26.2 | 22.0 | 27.3 |  |

**Table 5:** post COVID-19 complications of the three waves.

| Trait | Total patient | First wave | Second wave | Third wave | P value |
| --- | --- | --- | --- | --- | --- |
|  | 600 | 200 | 200 | 200 |  |
| Age, Mean<br>(SD) | 45.2(15.8) | 45.5(16.3) | 43.6(14.3) | 46.5(16.5) | 0.19 |
| Sex, Male n<br>(%) | 337(56.2) | 123(61.5) | 97(48.5) | 117(58.5) | 0.08 |
| Post COVID-19 conditions |  |  |  |  |  |
| Number of post-covid conditions |  |  |  |  | 0.24 |
| None | 40(6.7) | 17(8.5) | 12(6) | 11(5.5) |  |
| Single | 93(15.5) | 38(19) | 30(15.1) | 25(12.5) |  |
| Multiple | 466(77.7) | 145(72.5) | 157(78.9) | 164(82) |  |
| Symptoms |  |  |  |  |  |
| Feverish<br>feeling | 16(2.7) | 6(3) | 4(2) | 6(3) | 0.7 |
| Fatigue | 421(70.2) | 126(63) | 144(72) | 151(75.5) | 0.03 |
| Cough | 127(21.2) | 35(17.6) | 58(29) | 34(17) | 0.004 |
| Respiratory<br>distress | 40(6.7) | 14(7) | 15(7.5) | 11(5.5) | 0.7 |
| Hoarseness of<br>voice | 16(2.7) | 9(4.5) | 2(1) | 5(2.5) | 0.09 |
| Chest pain | 142(23.7) | 44(22.1) | 40(20.1) | 58(29) | 0.09 |
| Anorexia | 135(22.5) | 40(20) | 39(19.5) | 56(28) | 0.07 |
| Anosmia | 16(2.7) | 7(3.5) | 4(2) | 5(2.5) | 0.6 |
| Headache | 146(24.3) | 46(23) | 49(24.5) | 51(25.5) | 0.08 |
| Memory disturbance | 152(25.4) | 46(23) | 64(32) | 42(21.1) | 0.03 |
| Insomnia | 73(12.2) | 20(10) | 29(14.5) | 24(11) | 0.38 |
| Hypersomnia | 4(0.7) | 2(1) | 2(1) |  | 0.36 |
| Sleep pattern alteration | 6(1) | 2(1) | 2(1) | 2(1) | <0.99 |
| Arthralgia | 69(11.5) | 25(12.6) | 28(14) | 16(8) | 0.15 |
| Confusion | 29(4.8) | 5(2.5) | 12(6) | 12(6) | 0.18 |
| New onset hypertension | 15(2.5) | 2(1) | 7(3.5) | 6(3) | 0.23 |
| New onset Diabetes | 23(3.8) | 8(4) | 10(5) | 5(2.5) | 0.42 |
| Fatigue score, mean (SD) | 4.7(2.9) | 3.1(2.9) | 4.2(3.3) | 3.9(3.1) | 0.001 |
| Functional status, mean (SD) | 90.3(7.4) | 90.9(7.7) | 90(7.6) | 90(6.9) | 0.35 |
| Depression | 143(25.2) | 58(40.6) | 49(34.3) | 36(25.2) | 0.03 |

The daily COVID-19 infection rate was higher in the third wave and the duration of the second wave was short. The first wave lasted longer than the other wave, with an elevated and gradual infection rate decline.

The rate of testing was equal for all three waves. The positivity rate was higher during the third than in the first and second waves (Figure 2, Supplement 2).

**Figure 2:**
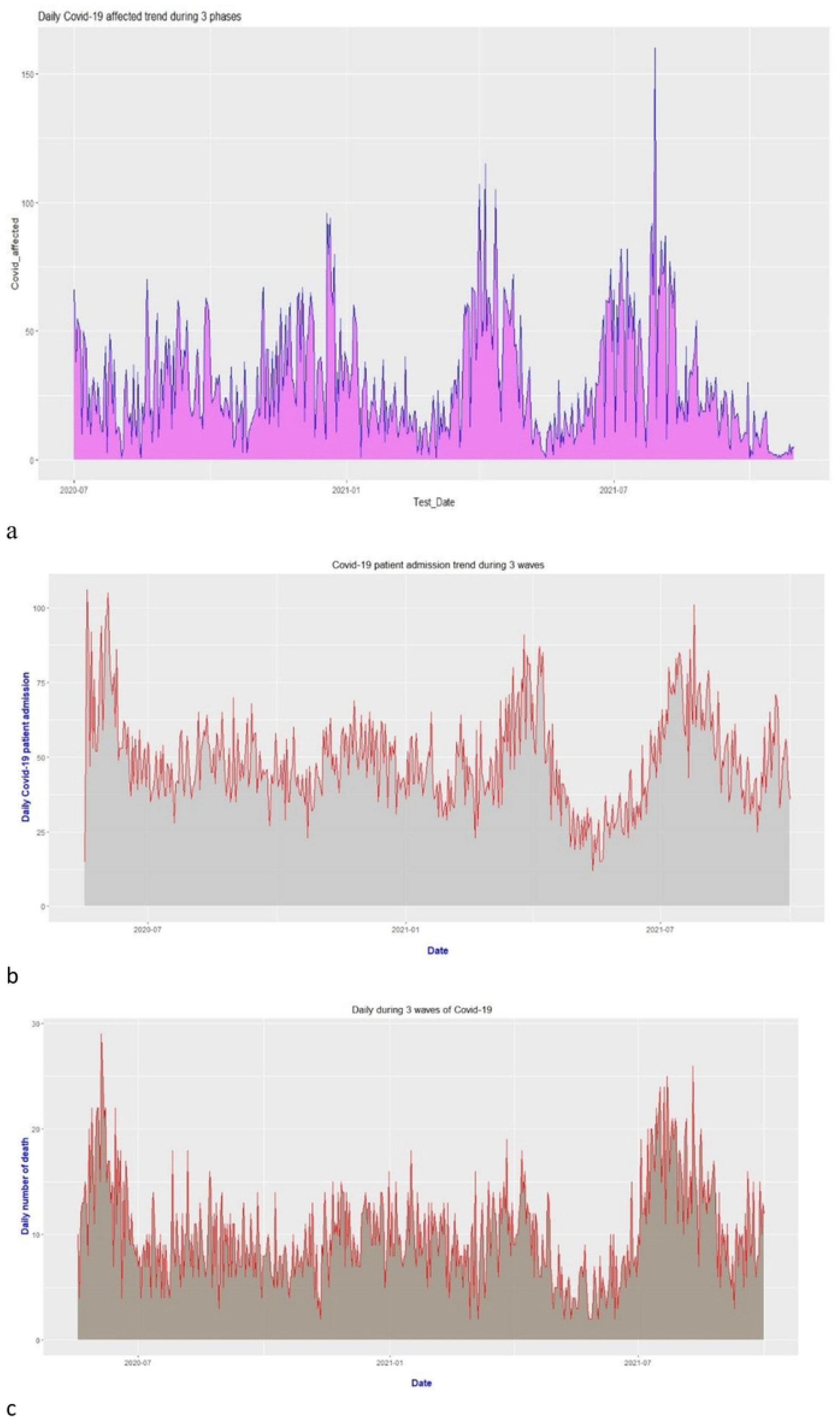
Epidemiologic trend of number of test positivity, hospital admission and death in three surges. a. Test positivity-The first surge remained longer and the second surge was narrowest. The daily number of positive cases was high in the second and third surge. b. Daily admission-Number of daily admissions was highest in the initial part of the first wave and third wave c. Daily death-Number of daily deaths was highest in the initial part of the first wave and third wave (The first, second, and third waves extend from April 2020 to January 2020, February 2021 to May 2021, and June 2021 to September 2021, respectively)

The percentage of patients that developed post-COVID complications was equal in all three waves. However, patients in the second wave developed more post-covid cough and memory disturbance depression, and patients in the third wave developed more fatigue than those in other waves. However, patients in the second wave had increased fatigue scores. The functional status of the patients was similar in the three waves. We found five symptom clusters. However, they did not differ among the three waves (Figure 3).

**Figure 3:**
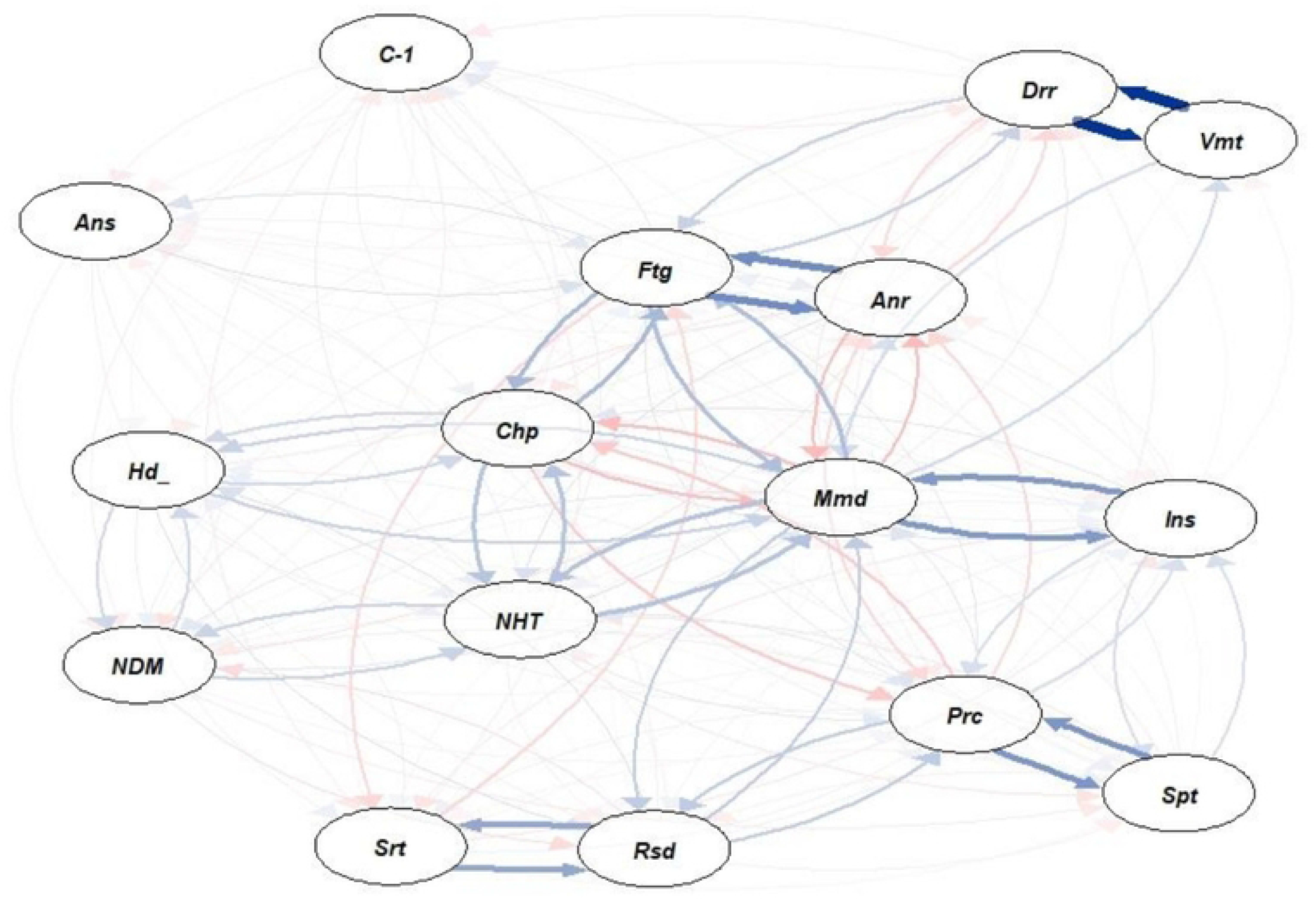
Symptom cluster in post COVID-state. There were five cluster of symptoms in the post-COVID state, 1-diarrhea and vomiting, 2-fatigue and anorexia, 3-memory loss and insomnia, 4-persistent cough and sputum, 5-sore throat and respiratory distress. We did the network analysis with tidy verse and q graph using R (v4.I. I). (cov-COVID-19, Ftg Fatigue, Ptc-persistent cough, Spt-sputum, Rsd-Respiratory distress, Srt-Sore throat, Chp-Chest pain, Drr-Diarrhea, Vmt-vomiting, Anr-Anorexia, Ans: Anosmia, Hdc-Headache, NHT: new onset hypertension, NDM: new onset diabetes, Mmd-Memory disturbance

## Discussion

This study compared the first, second, and third COVID-19 waves in terms of presentation, outcomes, and hospital epidemiological trends. The first wave lasted longer, and however, the second wave lasted for a short period. We found age and sex differences in the admission rates in the three waves. Cough, runny nose, respiratory distress, diarrhea, vomiting, and anosmia were the most common symptom in the three waves. The symptom cluster differed among the three waves. Disease severity, duration of hospital stay, and severity markers differed. We found disparities in the frequencies of the positivity test and death rates. The three waves had similar Post COVID complications.

This study was conducted at the tertiary center of Dhaka. The treatment facility differs from center to center and region to region; therefore, we cannot generalize the findings across the nation and globally.

During replication, a mutation in the genetic code of SARS-CoV-2 leads to the development of a new variant. The behaviors of variants, such as virulence and transmissibility, differ [28]. WHO named the variant to be monitored with the Greek alphabet and named from alpha to zeta [29]. In Bangladesh, until September 2021, alpha, beta, and delta variants were dominant at different times creating, three waves of COVID-19: the first wave was caused by alpha, the second wave was caused by beta, and the third wave caused by delta variants [18]. Epidemiological and disease characteristics differ in various studies [16,30,31].

In Bangladesh, the first wave lasted approximately 8 months, the second wave approximately 2 months, and the third wave approximately 3 months. This was due to the variations’ transmissibility; delta was 63–167% more transmissible than alpha [32]. Therefore, the most susceptible people became infected faster, replacing existing variants. Delta and beta variants spread quickly in Bangladesh, and delta quickly became the dominant variant. Omicron was 2.8 times more transmissible than Delta, thus replacing Delta in a short period [33].

In this study, we found a variation in the demographic patterns across the three variants.Young people were more affected in the initial wave, as evidenced by the high number of positive cases in the virology laboratory among the young. However, the elderly had a higher positivity rate. The proportion of older adult patients in the hospitalized patients was higher in the first and third waves than in the second wave. The admission rate among the attended patients in the outpatient department was higher in the second wave than in the first and third waves. Because of their increased mobility the younger age group became infected in large numbers. Immune senescence and various comorbidities could explain the older adults’ admission. The viral receptors ACE2 and CD26 are more expressed in senescent cells, which explains why older adults are more susceptible to infection [35]. The virus affinity for the receptor varies among the variants. This explains the age group variations among variants [36].

The overall number of admitted men was higher than women. However, the number of women admitted was higher in the second wave. Admission and death rates were higher among men. However,, women had a higher positivity rate than men, especially in the second and third waves. Men had higher rate of incidence of COVID-19 due to the altered immunologic response, associated comorbidity, hormonal differences, and smoking habits [35]. angiotensin-converting enzyme (ACE) receptor expression is higher in women because its genetic loci are on the X chromosome [37]. The *TMPRSS2* gene is located on chromosome 21q22.3[38]. Some variants have a higher affinity for TMPRSS than for ACE II. This might explain the gender differences in the expression of the various COVID-19 variants. Again, increased testosterone levels may increase the probability of microthrombi formation [39], which is the underlying pathophysiology of severe COVID-19.

The duration of Hospital admission was longer in the first wave, which was most likely due to the severity of the disease at the time of discharge. In the first wave, most patients were admitted because of isolation, and their release was determined by the time of PCR negativity. ICU admission was higher in the first wave due fear an unnecessary patient transfer to the ICU. The death rate was higher in the third wave than in other waves.

The different affinities of the viruses can explain the heterogeneous presentation and clustering of various variants to the receptor. The virus’s ability to escape immunity [40] can explain the mutant strain’s varied presentation and severity.

The three waves had slightly different post-COVID-19 complications. Further studies on its pathogenesis and immunological response are needed.

Because this was a single-center study, genotyping was either not performed or was impractical for all patients. This was a retrograde study and a review of the documents. Therefore, missing information was likely.

## Conclusion

This study revealed that during the COVID-19 waves, the presenting features, outcomes, and epidemiologic trends were different.

### Recommendation

In each of the three waves, COVID-19 occurred differently. A mutant strain of the SARS-CoV-2 was most likely the cause of these differences. Therefore, we recommend the following.

1. There should be genetic surveillance for the identification of the specific variant of interest.
2. At the onset of each wave, intensive epidemiologic surveillance should be conducted.
3. There should be diversity in each wave’s strategy for disease control planning, treatment, and hospital management.

## Data Availability

All data produced in the present study are available upon reasonable request to the authors

## Acknowledgments

We are grateful to Dhaka Medical College Hospital authority for giving us permission to access the hospital records; without their help it will be impossible to conduct this study.

## Supporting Information

Supplement 1: Patient selection and distribution of the patient for this cross-sectional study

Supplement 2: Difference in daily oxygen support, daily positive cases and daily test variability in three waves

